# The Swiss Integrated Care (INCA) Study: Description of a Novel Prospective Cohort of Patients and Caregivers in Reimbursed Informal Care

**DOI:** 10.64898/2026.07.14.26358029

**Authors:** Vasileios Nittas, Sandro Heiniger, Christina Haag, Anja Frei, Viktor von Wyl, Helena Aebersold, Maha Eid Madkour, Andreas Hellmann, Milo A. Puhan

**Affiliations:** Epidemiology, Biostatistics and Prevention Institute, University of Zurich, Zurich, Switzerland; MRC Cognition C Brain Sciences Unit, University of Cambridge, Cambridge, United Kingdom; Population Research Center, University of Zurich, Zurich, Switzerland; Institute for Implementation Science in Health Care, University of Zurich, Zurich, Switzerland; Entyre GmbH, Berlin, Germany

## Abstract

**Aims of the study:** This study aims to describe the methods, design, initial and planned implementation phases of the Swiss Integrated Care (INCA) cohort, as well as provide a descriptive analysis of cohort participants enrolled between July 2025 and March 2026. INCA is the first cohort in Switzerland to explore key aspects of reimbursed informal care as integrated in long-term care at home, closing a critical research gap. It was designed to fill current knowledge gaps and generate evidence on how reimbursed informal care is associated with the health and well-being of patients and caregivers.

**Methods:** INCA is a prospective, single-center cohort study with nationwide recruitment. Participation is open to adult patients and informal caregivers who provide paid informal care through home care agencies (Spitex organizations) in Switzerland. Eligible participants are enrolled consecutively. The cohort’s primary outcome is health-related quality of life of patients, assessed monthly through patient-reported outcome measures. Secondary outcomes include home care needs, including the overall health and well-being of patients (measured semi-annually), the type, amount, and quality of care (recorded daily), and caregiver burden and resilience (measured quarterly). Additional analysis will include structured medical data, extracted from patient-provided documents using Optical Character Recognition (OCR) technology and analysed using Large Language Models (LLM).

**Results:** Since recruiting started in July 2025, the cohort has enrolled 855 patients and 851 caregivers. Among patients, 53% are female, with a median age of 73 years. Caregivers are predominantly female (72%) with a median age of 56 years. Most patients experience impairments in physical functioning and participation in social roles. Among them, 85% require less than two hours of care per day, though care needs vary considerably. This is further reflected in the multi-attribute utility, where the overall PROMIS-Preference (PROPr) score is very low for most patients, with a median of 0.102, indicating a substantial need for medical assistance and care. With a median of 9 comorbidities, health-related quality of life is overall low for most patients. Cardiovascular and endocrine & metabolic diseases are amongst the most prevalent, affecting 69% and 65% of patients with available diagnoses (n=771). Certain diagnostic pairs occur more frequently than expected by chance, suggesting underlying links between disease categories.

**Conclusions:** INCA responds to a growing policy need for robust evidence on how new models of informal care are associated with the health and well-being of patients and their caregivers. Its longitudinal design, combining patient- and caregiver-reported data with medical records and innovative data extraction methods, will lay the groundwork for a better understanding of new informal care models in real-world settings. INCA’s findings are expected to have significant policy relevance and contribute to evidence-based policies on long-term care at home in Switzerland.

## INTRODUCTION

Informal care is typically delivered by relatives, friends, acquaintances, or other non-professional caregivers on a voluntary, non-contractual, and non-reimbursed basis [1]. It includes support with activities of daily living (e.g., mobility, personal hygiene, eating), and instrumental activities of daily living (e.g., care coordination, medication management) [1]. The Swiss Federal Office of Public Health defines informal care as the provision of support tasks (e.g., psychosocial support, household management) and care-related tasks, typically provided alongside professional home care services which cover clinical care needs [2]. It forms the backbone of care in Europe, where up to 80% of long-term care is estimated to be informal [3]. Amid rising shortages of trained healthcare staff, demographic aging, and a rising number of multi-morbid patients, the importance of informal care is expected to further grow and keep care-dependent patients longer at home [3], [4]. Yet, despite the broader societal value, the burden of informal care falls almost entirely on carers [5]. Evidence suggests that caregiver burden can lead to burnout, reduced quality of life, loss of income, economic vulnerability, social isolation, and deteriorating mental and physical health [3], [6]. Chronic pressure and stress, driven by the unpredictability of informal care and the lack of control, further increase the risk of negative outcomes [7]. To ensure that future long-term patients and their caregivers receive the care and support they need will require policy reform and innovation [8].

In Switzerland, informal care is a core pillar of home-based long-term care, with an estimated 600,000 individuals caring for relatives, friends or other acquaintances [2]. Several research projects and reports have highlighted the central role of informal caregivers in the Swiss healthcare system, highlighting key challenges such as caregiver burden, coordination with formal care, and the need for better system-level integration and support [9], [10], [11]. A particularly innovative support model - reimbursed informal care - emerged following a 2019 Swiss Federal Court decision, which ruled that untrained informal caregivers may receive payment for their services, provided they are employed by home care agencies (Spitex organizations) [12]. Spitex organizations are the core pillar of home care in Switzerland, providing professional nursing, daily life/task support, care consultations, and psychological support. Swiss home care is reimbursed through three tariffs. Tariff A includes consultation, assessment and coordination activities, Tariff B covers treatment care services (e.g., wound care), and Tariff C basic care needs (e.g., hygiene needs). Private as well as public Spitex organizations can formally employ informal caregivers and provide (a) support services such as nursing support and care training, and (b) reimbursement for delivered care through social insurance claims [1], [7].

This model is increasingly adopted across Switzerland, with the German-speaking regions being at the forefront, provided by both private and public Spitex agencies. A recent report by the Swiss Federal Office of Public Health suggests a substantial rise in uptake [12]. Between 2022 and 2024, the number of Spitex-employed informal caregivers is estimated to have increased approximately ninefold, and the reimbursed hours of care more than sixfold [12]. The report also projects a continued rise in the number of organizations employing such caregivers [12]. Caregiver reimbursement typically only covers care tasks, however, much of what caregivers deliver daily is considered support [13]. While support is essential and valuable, clearly separating it from care has become a key policy concern, aiming to prevent reimbursement misuse [12]. Reimbursed informal care aims to mitigate caregiver burden, increase caregiver wellbeing, and improve both the overall quality and conditions of informal care delivery. While this model is spreading rapidly across Switzerland, there is limited evidence of its impact on the health and well-being of patients and their caregivers [12]

To address this evidence gap, we have designed and launched the Swiss Integrated Care (INCA) cohort study. INCA aims to fill current knowledge gaps and generate empirical evidence on the health and well-being of patients and their caregivers under reimbursement contracts. The term “integrated care” refers to the integration of informal care within formal care structures. Here, we (a) describe INCA’s methods, design, initial implementation phase, (b) provide a descriptive analysis of cohort participants enrolled between July 2025 and December 2025, and (c) outline planned next steps.

## METHODS

### Study design

INCA is a prospective, single-center cohort study with nationwide recruitment, established in January 2025 for an initial duration of 10 years, and conducted at the University of Zurich (UZH) Epidemiology, Biostatistics and Prevention Institute (EBPI). INCA aims to examine how reimbursed informal care is associated with the health-related quality of life, health, functioning, and overall well-being of patients, as well as the burden and resilience of informal caregivers. INCA is an observational study and does not introduce a new model of care, nor does it modify existing informal care provision. The study has been approved by the Ethics Committee Zurich (Protocol number: 2024-01767). A protocol was registered with ISRCTN in February 2025 (ISRCTN16865563) [14].

### Population and setting

Participation is open to adult patients (primary participants) and their informal caregivers who provide paid informal care through a Spitex organization in Switzerland. Recruitment began in July 2025 with our first Spitex partner Pflegewegweiser, using their patient collective. Pflegewegweiser is among the largest reimbursed informal care providers in German-speaking Switzerland, active in 16 approved cantons and headquartered in St. Gallen [15]. INCA is designed as an open, ongoing cohort, and additional Spitex organizations are expected to contribute over time.

Currently, to be eligible, patients must be (a) 18 years of age or older, (b) have a nursing care prescription, (c) have adequate knowledge of the German language, and (d) be cognitively able to provide informed consent. Patients are the primary cohort participants. Once a patient agrees to participate, their caregiver is also invited to participate – through a separate consent procedure. Informal caregivers of participating patients must also be (a) 18 years of age or older, (b) have adequate knowledge of the German language, and (c) be cognitively able to provide informed consent. The German language requirement will be dropped for future recruitment of patients and caregivers in other parts of Switzerland, as soon as additional Spitex organizations join INCA.

Eligible patients and informal caregivers of collaborating Spitex organizations are enrolled with a consecutive sampling approach. Recruitment occurs via two parallel-running streams: (a) online, and (b) printed. In the online stream, patients and caregivers can participate in the cohort through a digital informed consent procedure. A study flyer with two QR codes (separate for patients and caregivers) is sent to all potential participants via E-Mail through Pflegewegweiser. Through the QR code, patients/caregivers can access the informed consent form (ICF) on a secure online data management platform (REDCap). To provide consent, patients must provide/confirm their full name, date of birth, email and residential postcode. Caregivers provide their full name, date of birth, email address. as well as the name of the person they are caring for. The printed stream requires physical signatures on paper-based ICFs with pre-filled data (full name, date of birth, email and residential postcodes). Those are handed out by trained nurses during initial or follow-up (one week after initial visit) appointments together with printed study flyers. Our current informed consent approach deviates from our registered protocol, as we additionally added printed ICFs [14]. The rationale behind that change was practical, as most patients and caregivers found it easier to sign on paper.

## Measures

### Primary Outcomes

#### Health-related Quality of Life

INCA’s initial primary outcome is health-related quality of life (HRQoL) of patients in reimbursed informal care, routinely collected by Pflegewegweiser using the PROMIS-29 *(Patient-Reported Outcomes Measurement Information System)* questionnaire [16], [17]. It contains 29 items covering seven health domains: physical function, anxiety, depression, fatigue, sleep disturbance, social role satisfaction, and pain interference, with an additional item assessing pain intensity. It has been extensively validated for its reliability and validity in evaluating health outcomes in home care settings and across various populations, showing excellent internal consistency (Cronbach’s alpha > 0.90) and test-retest reliability (ICC > 0.8), along with strong construct validity confirmed by its correlation with established health measures [18]. For INCA, we used the officially validated German-language version, which is made available through the PROMIS National Center at Charité, Berlin, under the corresponding arrangement, and applied in accordance with the applicable terms of use. PROMIS-29 data are collected monthly directly from patients and their caregivers via online questionnaires.

To quantify overall health-related quality of life (HRQoL) in the participants enrolled to date, we calculated the PROMIS-Preference (PROPr) score, a societal preference-based utility measure derived from seven PROMIS domains: cognitive abilities, depression, fatigue, pain interference, physical function, sleep disturbance, and ability to participate in social roles and activities [19]. The PROPr scoring algorithm applies single-attribute functions to transform domain-specific PROMIS responses into utilities, which are then combined using a multiplicative multi-attribute function to generate a summary score on a scale anchored at 0, representing death, and 1, representing full health. Because the PROMIS-29 version used in this study differs slightly from the original item set on which PROPr was developed, cognitive function was approximated using the available questionnaire items. Previous studies have shown moderate to strong correlations between PROPr and other HRQoL instruments, including EQ-5D measures, supporting its construct validity across different settings [20], [21], [22].

### Secondary outcomes

#### Home care needs

The overall health status, functioning, needs, and preferences of patients is assessed semi-annually using the *interRAI HCSchweiz (International Resident Assessment Instrument Home Care Switzerland)* questionnaire, designed as a person-specific needs assessment for the Swiss home care context and primarily for adult populations receiving home care and support from a home care organization [23]. The interRAI covers several key areas, including administrative data, admission history, cognitive abilities, communication and vision, mood and behavior, psychosocial well-being, physical functioning, continence, medical diagnoses and general health. *InterRAI* is collected by a responsible registered nurse during routine care planning processes and re-evaluated regularly to ensure continuity and adaptive care management.

#### Volume of prescribed care

The physician’s care prescription forms the basis of each care setting and specifies in detail the care tasks for which a recipient has been referred. For every prescribed task, the frequency and estimated duration are documented. Using this information, it is possible to calculate the expected volume of care in hours per month. The resulting measure provides an estimate of the total amount of informal care that a recipient is entitled to receive and serves as an important indicator of the individual’s care needs and required support intensity.

#### Daily care

The type, amount, and quality of provided informal care is assessed through data retrieved from the daily care documentation of caregivers. Once a day, caregivers are asked to document (free text or recording) a randomly selected care task, in addition to rating (a) their sense of security while performing the task, (b) the well-being of the person they are caring for, and (c) their own current emotional state.

#### Caregiver burden and resilience

Caregiver burden and resilience are captured quarterly with the FARBE (“Fragebogen zur Angehörigenresilienz und -belastung”) questionnaire, designed to assess the resilience (e.g., adaptability, coping strategies, emotional support, and optimism) and burden (e.g., physical strain, emotional stress, financial insecurity, and social impact) of family members facing significant health challenges or stressful life circumstances [24]. FARBE is completed by informal caregivers via email and on a voluntary basis. This quarterly assessment aims to complement the daily care documentation by capturing longitudinal changes in caregiver experience and perceived care quality. FARBE data is collected by the project coordination team via secure online submission.

#### Unstructured medical data

We continuously collect additional medical information from patient-provided documents, including medical reports, discharge summaries, medication lists, and diagnosis records. These predominantly unstructured documents are digitized using Optical Character Recognition (OCR) technology to convert their contents into machine-readable text. A large language model (LLM) is subsequently applied to identify, classify, and extract clinically relevant diagnostic and medication-related information. Extracted diagnoses are mapped to the ICD-10 classification system, comprising 22 main chapters with progressively more specific subchapters. As the present study focuses on patients’ comorbidity profiles, only chronic diseases were retained for analysis, whereas acute conditions were excluded.

The complete extraction workflow underwent systematic internal validation and quality assessment by medical experts, who compared the model-generated outputs with the original documents to assess their accuracy, consistency, and reliability. The validation demonstrated that the approach is feasible and sufficiently reliable for application in the present study. A detailed description of the extraction pipeline and the results of the validation are provided in the Appendix 1.

### Data collection

All primary and secondary outcomes are routinely collected by the home care agency Pflegewegweiser and are part of the reimbursement agreement. Raw data for all participants agreeing to participate in the INCA cohort will be made accessible by Pflegewegweiser to the EBPI team on a regular basis. In addition to these routine measures, we will periodically introduce additional surveys. This approach leverages existing data while remaining open to incorporating additional assessments as needed, ensuring flexibility and responsiveness to emerging evidence. A visual overview of routinely assessed measures is provided in Figure 1.

**Figure 1.**
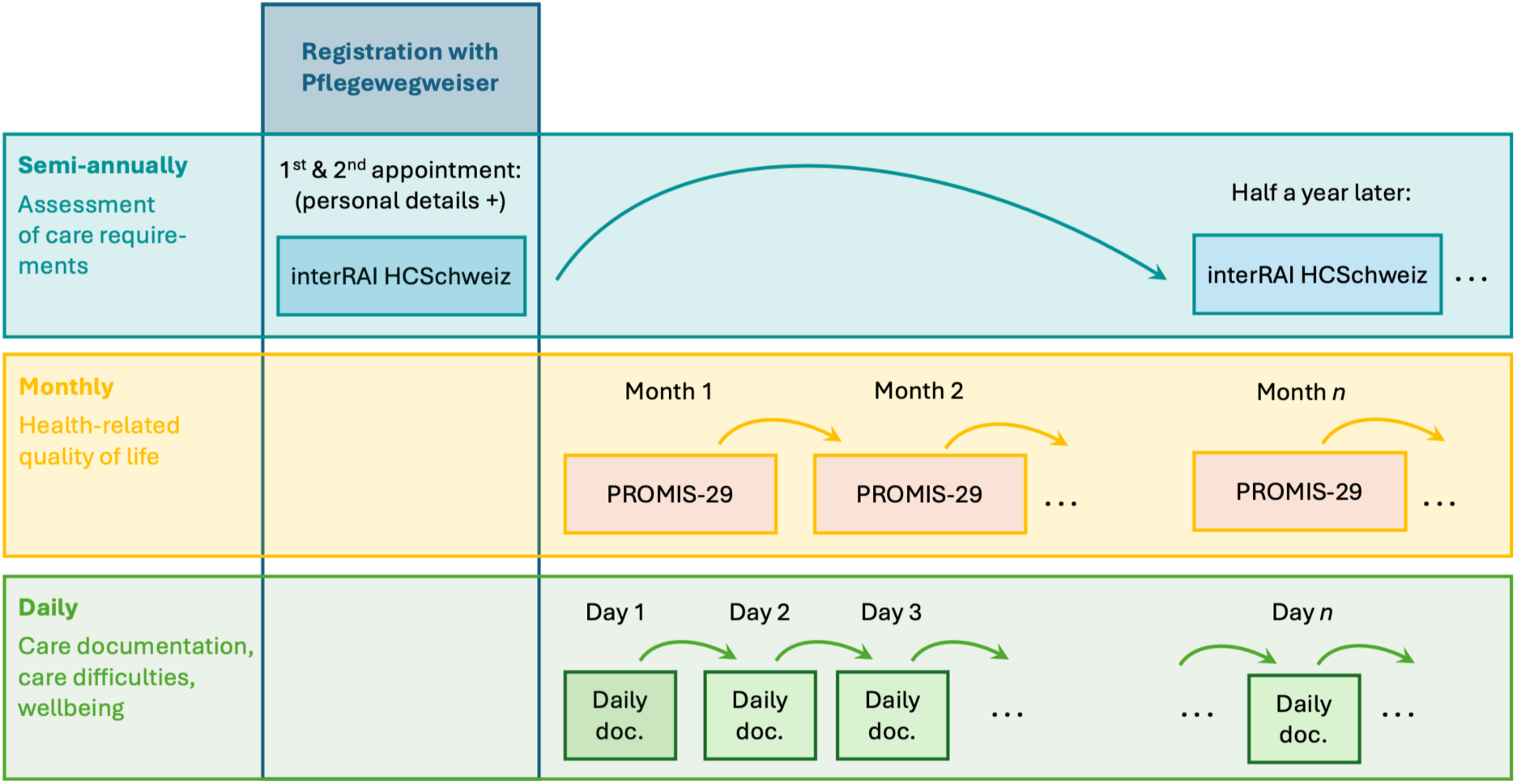
Routinely collected INCA cohort data.

### Statistical analysis

#### Analyses Conducted in the Present Study

Statistical analyses in this study were primarily descriptive and aimed at characterizing the current population enrolled in the INCA study. Cohort demographics of the INCA sample were summarized and compared with those of the overall Pflegewegweiser population to assess the representativeness of the study sample. To further describe the INCA cohort, descriptive statistics were calculated for estimated patient care needs, including measures of central tendency and dispersion. In addition, the distribution of caregiving tasks was analyzed by task category and tariff type to provide a more detailed overview of the nature and complexity of the care activities performed within the study population.

We further characterize the INCA population using the recorded diagnostic information. First, we describe the prevalence of conditions and diagnoses across ICD-10 chapters to provide an overview of the clinical profile of the sample. To explore diagnostic patterns in greater detail, we then assess comorbidity among patients. Specifically, we compare the observed co-occurrence of diagnoses across ICD-10 chapters with the prevalence expected under the assumption of random overlap between conditions. Under this assumption, the expected prevalence is calculated as the product of the marginal prevalence rates of the respective chapters. This approach allows us to identify diagnosis pairs that occur more frequently than would be expected by chance alone, thereby highlighting clinically relevant patterns of multimorbidity within the cohort.

#### Outline of Future Analytic Approaches

Future analyses will examine the type, amount, and quality of informal care provided to study participants, with particular attention to longitudinal patient trajectories over time. These analyses aim to identify factors associated with care recipients’ health-related quality of life, physical and mental health, daily functioning, and overall well-being. By modelling changes and developments across repeated observations, the study seeks to provide a comprehensive understanding of how different aspects of reimbursed informal care contribute to individual and collective health outcomes.

For each specific research question and analytic focus, a detailed and fully specified analysis plan will be presented in the respective subsequent publications, reflecting the evolving scope of the study and the accumulating data.

#### Open Science and Data Availability

In line with open science principles, the analysis files, including the complete analysis code and detailed information on the software environment, libraries, and R packages used, are publicly available via the Harvard Dataverse [27]. To ensure compliance with ethical, legal, and contractual obligations, only data that can be publicly shared are included. Sensitive patient-level data, as well as data owned by partner commercial organizations, are excluded and cannot be made publicly available.

## RESULTS

### Study status

Since recruiting was launched in July 2025, the INCA cohort has enrolled 855 patients and 851 caregivers (status: April 16^th^, 2026). Recruitment is ongoing with an average of 23 weekly patients and 23 weekly caregiver enrolments, respectively. Below, we provide a first description of the current cohort. Figure 2 shows participant selection based on STROBE [28].

**Figure 2.**
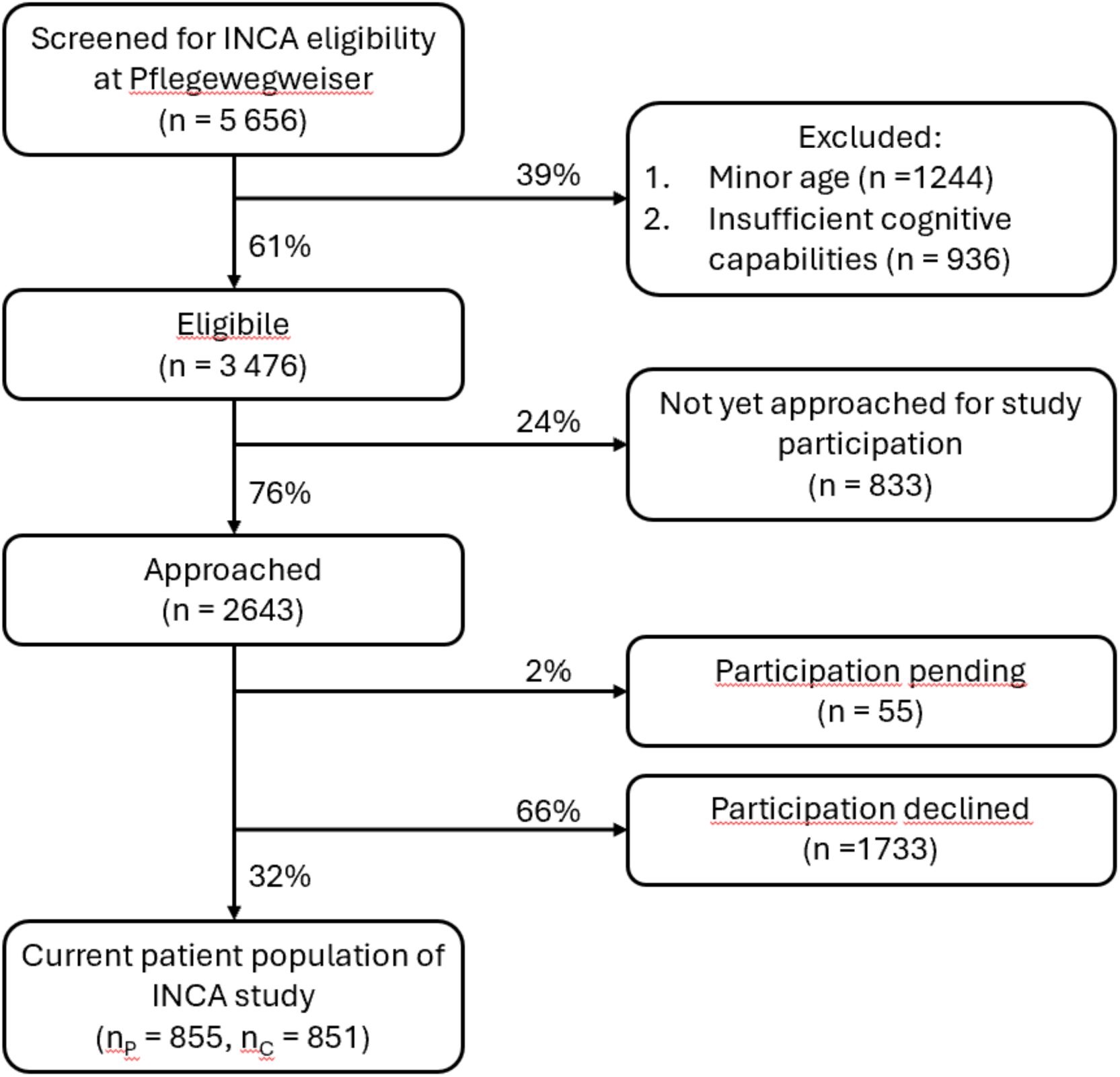
Strobe chart on study sample and participation. *Notes: n = number of patient/caregiver relationships; np=number of patients; nc= number of caregivers. A mutual exclusion restriction applies when the patient is excluded, which also leads to exclusion of the caregiver; however, exclusion of a caregiver does not affect the patient’s eligibility for the study population. Therefore, the counts in the respective categories do not need to sum correctly*.

### Demographics

Descriptive statistics on the demographics of patients and caregivers are provided in Table 1 and reveal distinct patterns in sex, age, nationality, and professional background. Among patients, the sex ratio is nearly balanced, with 53% identified as female. In contrast, the caregiver population is predominantly female, comprising 72% of all caregivers. Most patients receiving care are elderly, with a median age of 73 years (IQR: 23, 5%/95%-quantile: 29/92), although the data include younger patients across a wide age range. Caregivers are typically middle-aged, with a median age of 56 years (IQR: 25, 5%/95%-quantile: 25/79); however, substantial variation exists, with caregivers spanning from very young adults to older individuals. Most caregivers are Swiss nationals (70%). More than half of them are not employed elsewhere, while 44% provide informal care part-time alongside other employment. Only a small proportion of caregivers (13%) possess a professional qualification in care, enabling them to perform more complex Tariff B tasks. The right side of Table 1 shows the overall shares in the total population registered with Pflegewegweiser, and the demographics observed in the INCA cohort closely mirror this population.

**Table 1:**
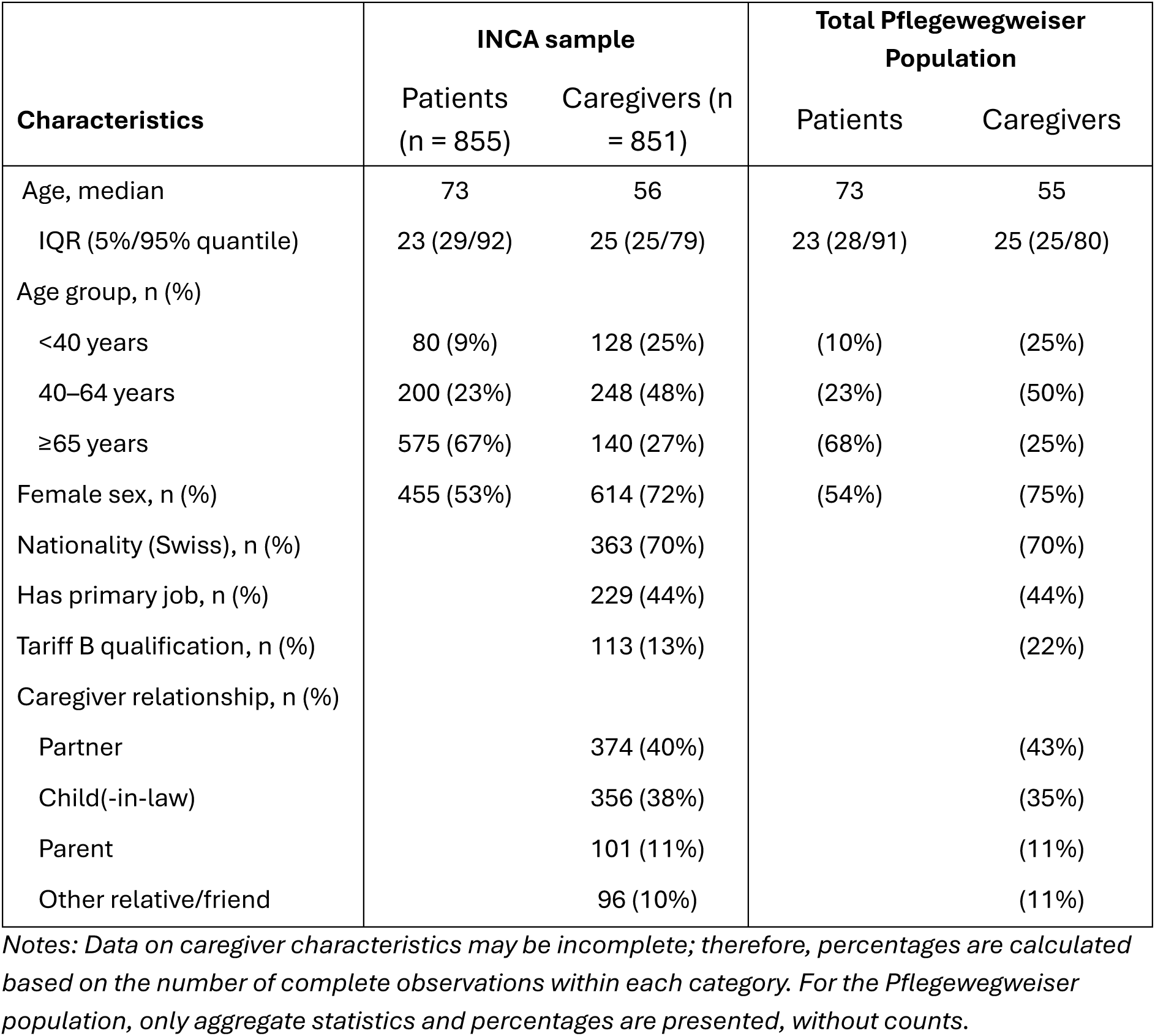
Cohort demographics.

| Characteristics | INCA sample |  | Total Pflegewegweiser Population |  |
| --- | --- | --- | --- | --- |
|  | Patients<br>(n = 855) | Caregivers (n<br>= 851) | Patients | Caregivers |
| Age, median | 73 | 56 | 73 | 55 |
| IQR (5%/95% quantile) | 23 (29/92) | 25 (25/79) | 23 (28/91) | 25 (25/80) |
| Age group, n (%) |  |  |  |  |
| <40 years | 80 (9%) | 128 (25%) | (10%) | (25%) |
| 40–64 years | 200 (23%) | 248 (48%) | (23%) | (50%) |
| ≥65 years | 575 (67%) | 140 (27%) | (68%) | (25%) |
| Female sex, n (%) | 455 (53%) | 614 (72%) | (54%) | (75%) |
| Nationality (Swiss), n (%) |  | 363 (70%) |  | (70%) |
| Has primary job, n (%) |  | 229 (44%) |  | (44%) |
| Tariff B qualification, n (%) |  | 113 (13%) |  | (22%) |
| Caregiver relationship, n (%) |  |  |  |  |
| Partner |  | 374 (40%) |  | (43%) |
| Child(-in-law) |  | 356 (38%) |  | (35%) |
| Parent |  | 101 (11%) |  | (11%) |
| Other relative/friend |  | 96 (10%) |  | (11%) |

*Notes: Data on caregiver characteristics may be incomplete; therefore, percentages are calculated based on the number of complete observations within each category. For the Pflegewegweiser population, only aggregate statistics and percentages are presented, without counts.*

### Caregiver-Patient relationship

A large proportion of caregivers currently enrolled in the INCA cohort are partners of the patients, accounting for 40% of all caregivers. The second most common relationship is that of a child or child- in-law, which together represent 38% of caregivers, followed by parents (11%) and other relatives or friends (10%). This pattern largely explains the substantial age gap observed between patients and caregivers. In less frequent cases, caregivers are other relatives or even friends, highlighting that while most care is provided by close family members, the network of informal support can extend beyond immediate family.

### Patient care needs

The estimated monthly care is illustrated in Figure 2. Patients in the study population have an average estimated care need of 40.4 hours per month (median: 36 h, SD: 22.3 h), based on care prescriptions resulting from physicians’ assessments and evaluations. While most patients require a modest amount of care, the range of needs is considerable, with some requiring several hours of assistance daily. Care requirements did not differ notably by sex or by age group. It is important to note that informal care primarily covers basic support tasks that relatives can provide, often with limited formal training. In 16% of cases, a qualified nursing professional from another Spitex organization is additionally involved, allowing trained caregivers to perform more complex care tasks that require specialized education and expertise.

**Figure 2.**
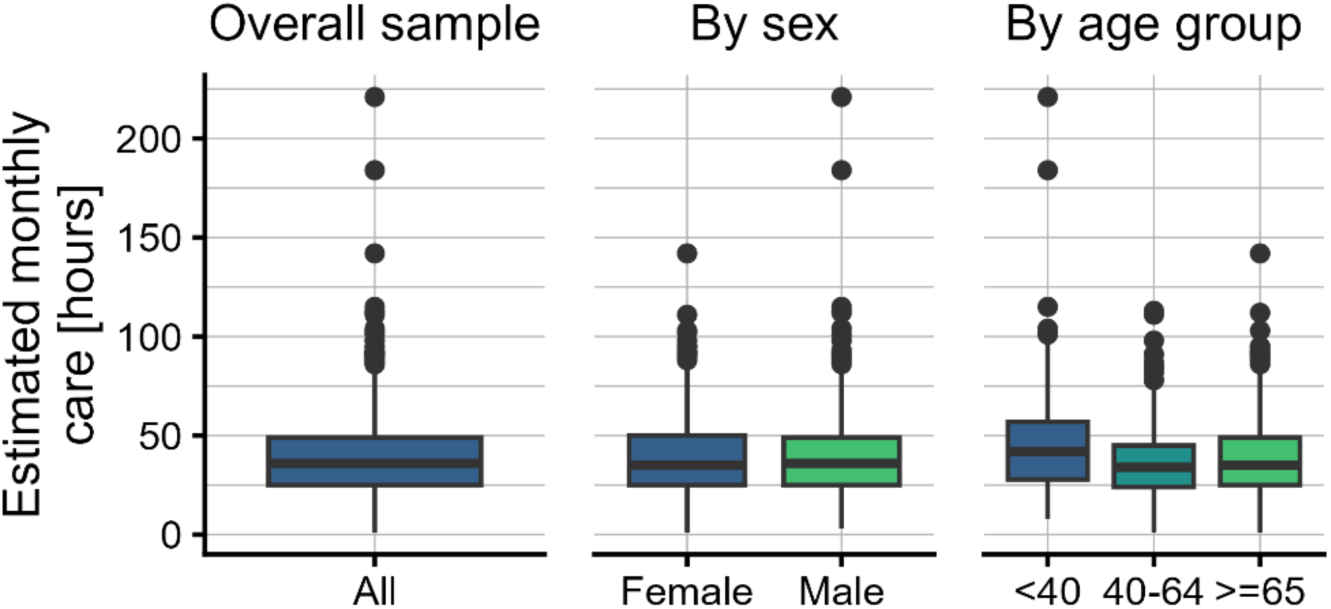
Estimated patient care needs based on care prescriptions. *Notes: Data are based on care prescriptions from 855 patients. Only each patient’s most recent prescription is included. The boxplot shows the median and interquartile range (IQR), with whiskers extending from the box to the smallest and largest values within 1.5 × IQR from Q1 and Q3, respectively*.

Patients in the study sample are mostly impaired in their physical functioning and in participation in social roles and activities. Moderate impairments were observed in pain interference, whereas cognitive function and sleep disturbance were less affected. This pattern aligns with the demographic composition of the study population. The multi-attribute utility, i.e., the overall PROPr score, is very low for most patients, reflecting a substantial need for medical assistance and care. Figure 3 displays the utility results by PROMIS domain and the derived PROPr score.

**Figure 3:**
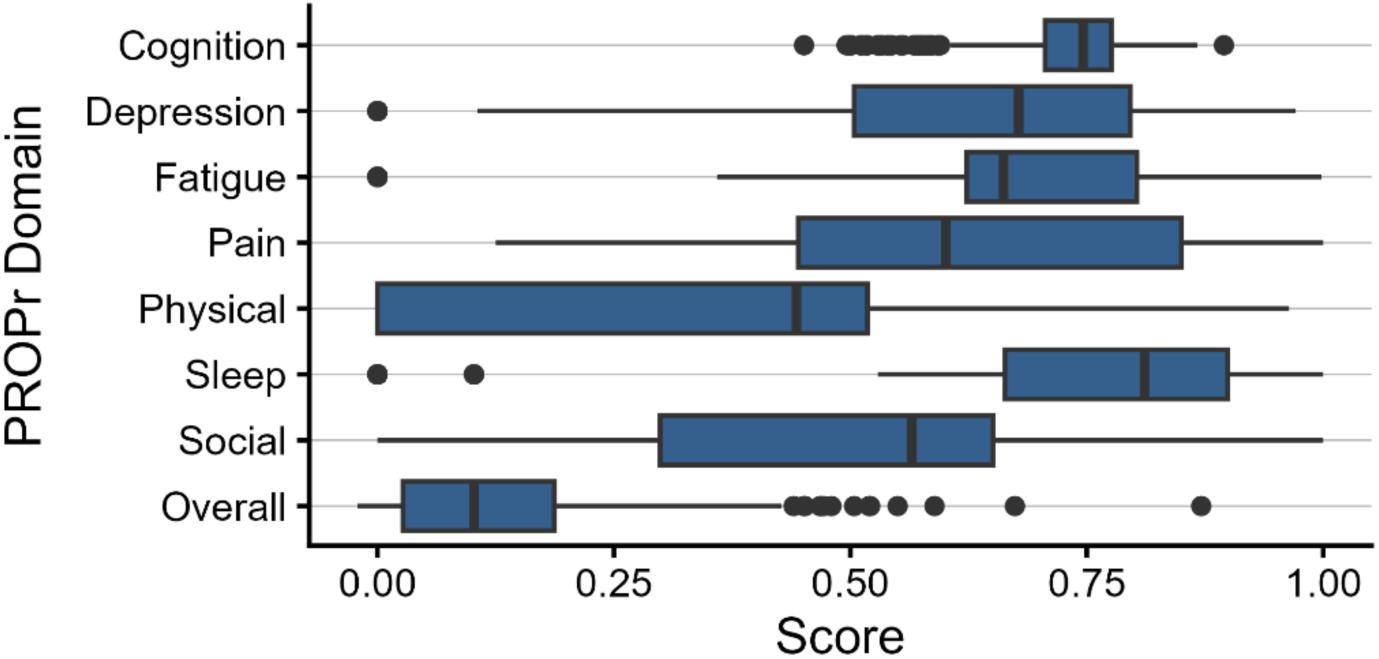
Overall and by-category PROPr scores among current cohort participants. *Notes: Responses from 507 patients; only the most recent response per patient is shown. The boxplot shows the median and interquartile range (IQR), with whiskers extend from the box to the smallest and largest values within 1.5 × IQR from Q1 and Q3, respectively. Higher scores in each PROMIS domain indicate better health status. The overall PROPr score is anchored on a scale from 0 to 1, where 0 represents a health state equivalent to death and 1 represents full health*.

The distribution of care tasks by tariff category in Table 2 further illustrates the predominance of basic support activities in the study population. Most observed care hours were allocated to tasks related to hygiene and comfort, accounting for 83’998 hours and representing the largest share of all documented activities. In contrast, tasks performed by caregivers with a Tariff B qualification accounted for only 2’032 hours, corresponding to approximately 2% of the total observed care hours. Tariff B caregivers are qualified to provide more specialized nursing and medical support tasks, which is reflected in the distribution of activities within this tariff category. The majority of Tariff B hours were associated with administering treatments and monitoring vital signs, indicating that these caregivers primarily contribute to more clinically complex and medically oriented aspects of care.

**Table 2:** Observed volume of care tasks per tariff and by task category in hours.

| Task category | Tariff C | Tariff B |
| --- | --- | --- |
| Bandages and assistive devices | - | 73 |
| Breathing | - | 80 |
| Excretion | 5'009 | 63 |
| Hygiene and comfort | 83'998 | 25 |
| Mental health services | - | 65 |
| Mobilization | 6'358 | - |
| Monitoring vital signs | - | 355 |
| Nutrition | 2'021 | 165 |
| Treatments | 120 | 1'206 |
| **Total** | **97'506** | **2'032** |

*Notes: Durations represent estimated standard task times rather than the actual time spent delivering care. Values are aggregated hours of care tasks by tariff category and task category. A dash (“-”) indicates that no tasks were recorded for the respective tariff within that category.*

### Patient diagnoses and multimorbidity

For 771 patients, representing 90.2% of the study sample, diagnosis information could be successfully extracted through natural language processing of medical documents (Figure 4). The median count of different diagnoses these patients have is 9 (IQR: 6), which indicates considerable variability in the complexity of health conditions within the study population. The most commonly observed ICD-10 chapter is ‘*diseases of the circulatory system’*, affecting 69% of the patients. This is followed closely by ‘*Endocrine, nutritional and metabolic diseases’*, which are present in 65% of the patients. These findings highlight the high prevalence of cardiovascular and metabolic health issues among the care recipients in the study.

**Figure 4:**
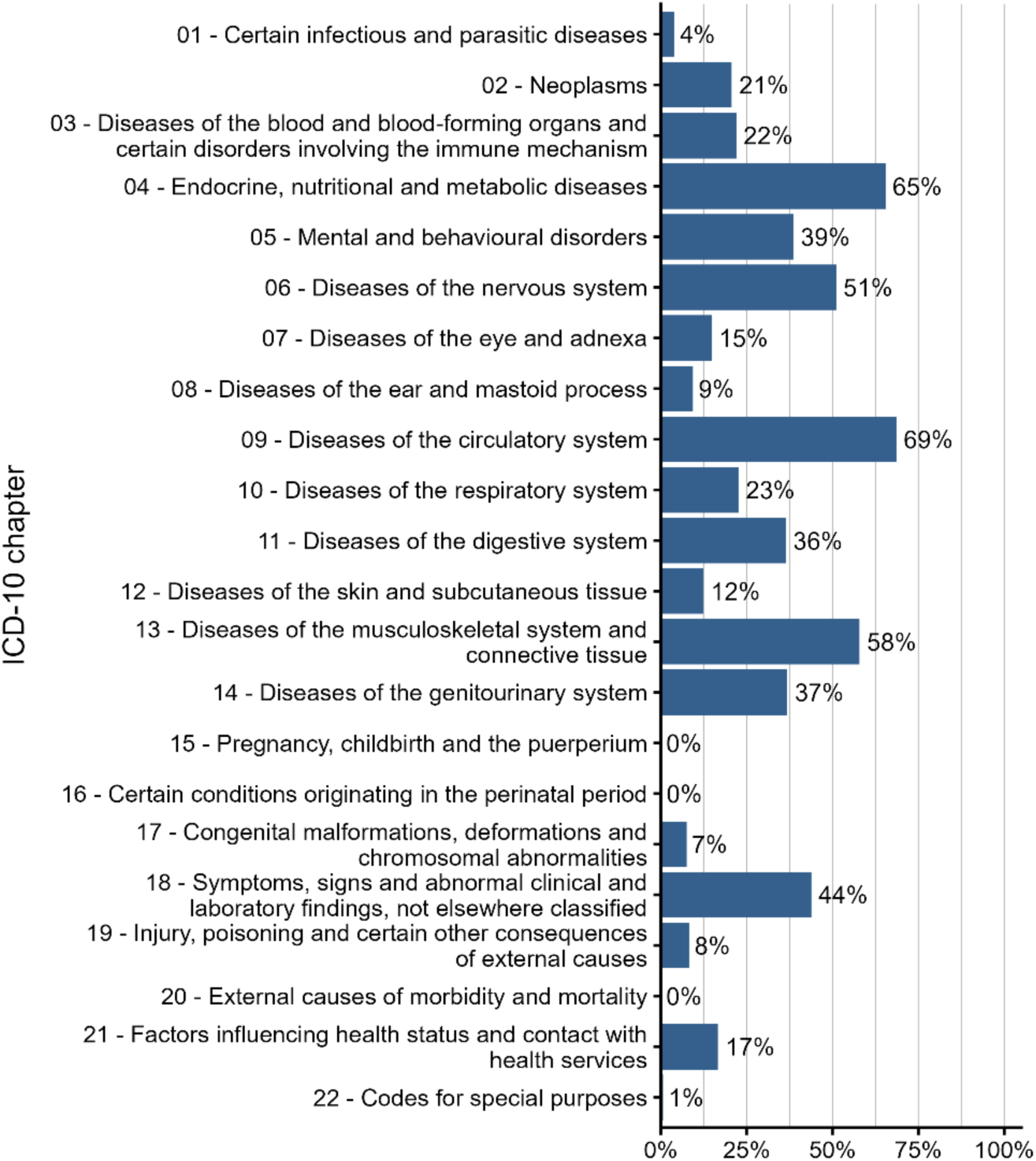
Prevalence of conditions and diagnoses by ICD-10 chapters of 726 patients.

The analysis of diagnosis combinations in Figure 5 reveals that certain pairs occur together more frequently than would be expected by chance. In particular, patients with a diagnosis related to the ‘*genitourinary system’* (ICD-10 chapter 14) often also have a diagnosis of a ‘*disease of the digestive system’* (11). Similarly, diagnoses in the ‘*circulatory system’* (09) frequently co-occur with ‘*endocrine, nutritional and metabolic diseases’* (04), but they very rarely coincide with ‘*diseases of the nervous system’* (06) or ‘*mental and behavioural disorders’* (05). Furthermore, we observe that ‘*diseases of the musculoskeletal system and connective tissue’* (13), one of the most prevalent diagnostic categories, seldom occur together with *‘neooplasms’* (02), ‘*diseases of the nervous system’* (06), or ‘diseases of the *circulatory system’* (09). These associations suggest underlying pathophysiological or age-related links between certain disease categories within the study population.

**Figure 5:**
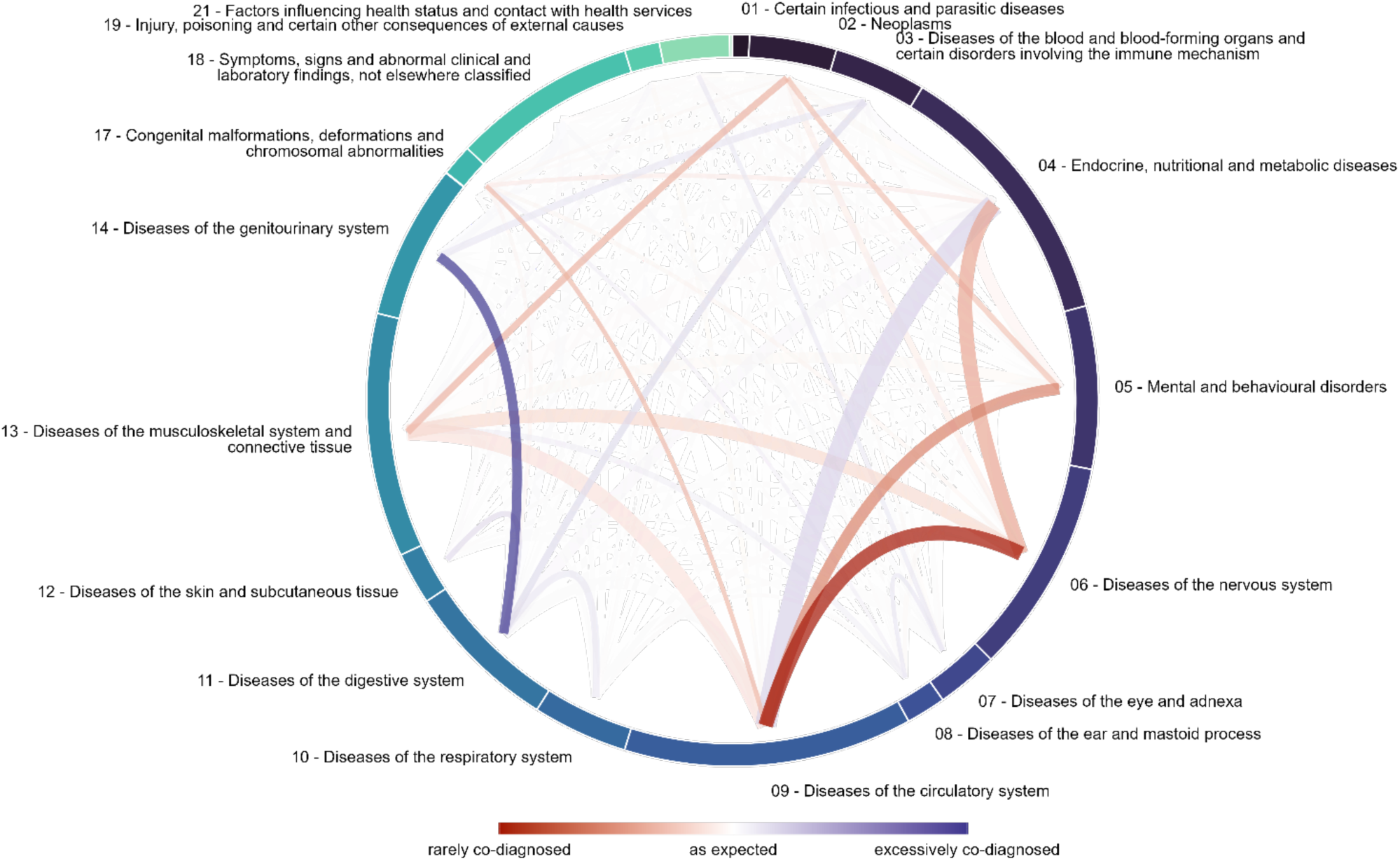
Co-occurrence of diagnoses compared to expected prevalence based on 726 patients.

## DISCUSSION

INCA is the first cohort in Switzerland that studies key aspects of reimbursed informal care, closing a critical evidence gap in home care research. By including patients and their caregivers across several regions in Switzerland, INCA responds to a growing policy need for robust evidence on how new models of informal care are associated with patient and caregiver health and wellbeing, as well as overall home care sustainability. Our cohort is also aligned with European calls for a stronger focus on the well-being of informal caregivers and a stronger linkage between formal and informal structures [21], [29]. Over time, INCA will generate data that could help establish benchmark trajectories for HRQoL and caregiver burden under reimbursed informal care and identifying predictors of outcome heterogeneity, informing quality improvement and innovation. INCA’s longitudinal outcome data (PROMIS-29, FARBE) and care process documentation will identify optimal implementation conditions for evidence-based workforce planning, balanced with safeguards for standardized training, burden monitoring, and quality assurance.

Our initial descriptive data highlight that INCA, through the close collaboration with one of the largest and fastest growing reimbursed informal care providers in Switzerland, successfully reaches a diverse patient population and their caregivers. Although participants vary in age and care intensity, our current cohort includes more older patients with multiple chronic conditions and mobility restrictions, as well as predominantly female caregivers. This is in line with previous research that shows that most home-dwelling elderly in Switzerland are multimorbid and with complex care needs, while most informal caregivers are middle-aged females [30], [31]. The observed empirical distribution of PROPr scores is consistent with findings from other comparable study populations [22], [32]. While these findings provide a first descriptive glimpse into our cohort, INCAs strength lies in its longitudinal approach and the findings that are yet to come. Ongoing Swiss projects, such as the SCOHPICA cohort of healthcare professionals and informal caregivers, also aim to longitudinally capture the needs and preferences of informal caregivers [9]. Similar to INCA, the SCOHPICA cohort consists predominantly of female and substantial proportion of working-age caregivers [9]. While INCA and SCOPHICA focus on different populations and regions, they highlight consistent caregiver profiles across Switzerland as well as the growing recognition of informal care as a key component of the Swiss healthcare system.

From a methodological perspective, INCA aims to demonstrate the feasibility of monitoring and evaluating new models of informal care at scale. Our approach improves data completeness and ecological validity while keeping the burden on elderly patients and their caregivers minimal [33]. INCA’s longitudinal design will allow a better understanding of care trajectories as well as the association of reimbursement with patient and caregiver health and well-being. From a policy perspective, INCA addresses a key gap in Swiss home care policy: the integration of patient and caregiver perspectives into outcome assessment and reimbursement decisions [12]. Additionally, the use of patient-reported outcomes (PROMIS-29) is aligned with international efforts toward person- centered care [34].

### Limitations

INCA has several limitations. First, current participation requires sufficient proficiency of German. This limits the inclusion of non-German speaking Swiss residents as well as non-German speaking migrant populations. Through the future inclusion of additional Spitex partners, we expect to overcome language barriers and reach the non-German speaking parts of Switzerland. Second, recruitment is currently limited to one study partner. Although the demographic characteristics of the INCA cohort closely mirror those of the overall Pflegewegweiser population, suggesting good internal representativeness, organization-specific practices might limit overall generalizability. We will overcome these limitations with the gradual inclusion of additional Spitex partners. Third, a substantial share of INCA is self-reported, which might be subject to biases. To mitigate this risk, we combine self-reported measures with routinely collected clinical assessments (interRAI), care prescriptions, and daily care documentation. Finally, INCA is an observational cohort without a control group. While its longitudinal design allows the analysis of within-cohort trajectories, it does not allow for any causal claims regarding the effects of reimbursed informal care on patient and caregiver health and well-being.

## CONCLUSION

INCA is a novel cohort study that aims to generate much-needed evidence on how reimbursed informal care is associated with the health and well-being of patients and informal caregivers in Switzerland. Its longitudinal design, combining patient- and caregiver-reported data with medical records and innovative data extraction methods, will lay the groundwork for a better understanding of new informal care models in real-world settings. In a rapidly changing care environment across Europe, our findings aim to provide policy- and practice-relevant insights into how reimbursed informal care affects patient outcomes, caregiver well-being, and the overall sustainability of home-based long-term care.

## Funding

Pflegewegweiser partially funded this study and supported participant recruitment and data provision. Beyond the contribution of A.H. as a co-author to the conception and design of the study (see Author Contributions), the funder had no role in data analysis, interpretation of results, or the decision to submit the manuscript for publication, which were carried out independently by the academic investigators.

## Conflicts of Interest

A.H. is Chief Medical Officer of Pflegewegweiser (the study funder) and of its parent company, Entyre GmbH. This role is disclosed as a potential conflict of interest. A.H. contributed to the conception and design of the study but was not involved in data analysis or interpretation, which were conducted independently by the academic investigators. All other authors declare no competing interests.

## Use of Generative AI

For this manuscript, generative AI (ChatGPT) was only used selectively for language editing support in order to improve readability and grammar.

## Notes

### Competing Interest Statement

The authors have declared no competing interest.

### Clinical Protocols

https://www.isrctn.com/ISRCTN16865563?q=ISRCTN16865563&filters=&sort=&offset=1&totalResults=1&page=1&pageSize=10

### Author Declarations

The Ethics Committee pf the Canton of Zurich gave ethical approval for thbis work (Protocol number: 2024-01767).

## References

1. [1] V. ZIgante, ‘Informal care in Europe Exploring formalisation, availability and quality’, 2018.

[2] BAG, ‘Betreuende und pflegende Angehörige’, https://www.bag.admin.ch/de/betreuende-und-pflegende-angehoerige.

[3] V. Zigante, ‘Informal care in Europe Exploring Formalisation, Availability and Quality’, 2018.

[4] L. Escasain et al., ‘Informal caregivers’ health: a literature review’, 2023, doi: 10.16908/issn.1660-7104/348.

[5] D. Bouget, S. Spasova, and B. Vanhercke, ‘Work-life balance measures for persons of working age with dependent relatives in Europe’, 2016.

[6] C. Y. Chiao, H. S. Wu, and C. Y. Hsiao, ‘Caregiver burden for informal caregivers of patients with dementia: A systematic review’, Sep. 01, 2015. doi: 10.1111/inr.12194.

[7] P. P. Vitaliano, J. Zhang, and J. M. Scanlan, ‘Is Caregiving Hazardous to One’s Physical Health? A Meta-Analysis’, Nov. 2003. doi: 10.1037/0033-2909.129.6.946.

[8] C. Lussi, J. Bickenbach, R. Halvorsen, and C. Sabariego, ‘Trends over the past 15 years in long-term care in Switzerland: a comparison with Germany, Italy, Norway, and the United Kingdom’, BMC Geriatr., vol. 24, no. 1, Dec. 2024, doi: 10.1186/s12877-024-05195-8.

[9] SCOHPICA, ‘April 23 2026 – SCOHPICA-Informal Caregivers: First Results from the 2025 Data Collection’, https://scohpica.ch/de/april-23-2026-scohpica-informal-caregivers-results-2025/.

[10] Berner Fachhochschule, ‘Projekt: Betreuende Angehörige’, https://www.bfh.ch/de/forschung/forschungsprojekte/2025-262-151-794/?utm_source=chatgpt.com.

[11] I. Peytremann-Bridevaux et al., ‘Protocol for the Swiss COhort of Healthcare Professionals and Informal CAregivers (SCOHPICA): Professional trajectories, intention to stay in or leave the job and well-being of healthcare professionals’, PLoS One, vol. 19, no. 8, p. e0309665, Aug. 2024, doi: 10.1371/journal.pone.0309665.

[12] Der Bundesrat, ‘Pflegeleistungen von Angehörigen im Rahmen der obligatorischen Krankenpflegeversicherung’, 2025.

[13] OECD, ‘Who Cares? Attracting and Retaining Care Workers for the Elderly’, 2020.

[14] M. Puhan, V. von Wyl, A. Frei, and V. Nittas, ‘A Swiss study on informal care patients and their caregivers’, 2025, London, UK. doi: 10.1186/ISRCTN16865563.

[15] Entyre GmBH, ‘Pflegewegweiser - Standorte’, https://pflegewegweiser.ch/standorte/.

[16] S. A. Rimehaug et al., ‘Measurement properties of the PROMIS-29 profile v2.1 in a Norwegian rehabilitation context’, J. Patient. Rep. Outcomes, vol. 9, no. 1, Dec. 2025, doi: 10.1186/s41687-025-00929-7.

[17] E. B. M. Elsman, L. D. Roorda, N. Smidt, H. C. W. de Vet, and C. B. Terwee, ‘Measurement properties of the Dutch PROMIS-29 v2.1 profile in people with and without chronic conditions’, Quality of Life Research, vol. 31, no. 12, pp. 3447–3458, Dec. 2022, doi: 10.1007/s11136-022-03171-6.

[18] M. C. Hwang, C. Bell, Y. Farran, A. Ogdie, C. Green, and J. Reveille, ‘Reliability and validity of the PROMIS-29 health profile in Ankylosing Spondylitis patients: A cross-sectional study’, Medicine (United States), vol. 103, no. 9, p. E37251, Mar. 2024, doi: 10.1097/MD.0000000000037251.

[19] B. Dewitt, H. Jalal, and J. Hanmer, ‘Computing PROPr Utility Scores for PROMIS® Profile Instruments’, Value in Health, vol. 23, no. 3, pp. 370–378, Mar. 2020, doi: 10.1016/j.jval.2019.09.2752.

[20] B. J. Mulhern et al., ‘Understanding the measurement relationship between EQ-5D-5L, PROMIS-29 and PROPr’, Quality of Life Research, vol. 32, no. 11, pp. 3147–3160, Nov. 2023, doi: 10.1007/s11136-023-03462-6.

[21] F. Colombo, A. Llena-Nozal, J. Mercier, and F. Tjadens, Help Wanted? Providing and pazing for long-term care. in OECD Health Policy Studies. OECD, 2011. doi: 10.1787/9789264097759-en.

[22] C. P. Klapproth, F. Fischer, M. Merbach, M. Rose, and A. Obbarius, ‘Psychometric properties of the PROMIS Preference score (PROPr) in patients with rheumatological and psychosomatic conditions’, BMC Rheumatol., vol. 6, no. 1, Dec. 2022, doi: 10.1186/s41927-022-00245-3.

[23] Spitex Schweiz ‘interRAI HomeCare Schweiz’, https://www.spitex-instrumente.ch/bedarfsabklaerung/interrai-instrumente/interrai-homecare-schweiz.

[24] A. Lübben, L. Peters, M. Przysucha, and A. Büscher, ‘Einfluss von Faktoren auf die Resilienz und Belastung pflegender Angehöriger (FARBE) – Fragenbogen zur Angehörigen-resilienz und - belastung’, Pravention und Gesundheitsforderung, vol. 19, no. 4, pp. 590–595, Nov. 2024, doi: 10.1007/s11553-023-01076-x.

[25] B. Dewitt et al., ‘Estimation of a Preference-Based Summary Score for the Patient-Reported Outcomes Measurement Information System: The PROMIS®-Preference (PROPr) Scoring System’, Medical Decision Making, vol. 38, no. 6, pp. 683–698, Aug. 2018, doi: 10.1177/0272989X18776637.

[26] X. Tang et al., ‘Mapping and Linking between the EQ-5D-5L and the PROMIS Domains in the United States’, Medical Decision Making, vol. 45, no. 6, pp. 740–752, Aug. 2025, doi: 10.1177/0272989X251340990.

[27] S. Heiniger, ‘Description of a Novel Prospective Cohort of Patients and Caregivers in Reimbursed Informal Care’, 2026.

[28] E. von Elm, D. G. Altman, M. Egger, S. J. Pocock, P. C. Gøtzsche, and J. P. Vandenbroucke, ‘Strengthening the reporting of observational studies in epidemiology (STROBE) statement: guidelines for reporting observational studies’, BMJ, vol. 335, no. 7624, pp. 806–808, Oct. 2007, doi: 10.1136/bmj.39335.541782.AD.

[29] E. Verbakel, S. Tamlagsronning, L. Winstone, E. L. Fjaer, and T. A. Eikemo, ‘Informal care in Europe: Findings from the European Social Survey (2014) special module on the social determinants of health’, 2017, Oxford University Press. doi: 10.1093/eurpub/ckw229.

[30] A. Chiolero, V. Burato Guttierrez, F. Clausen, and A. Chiolero, ‘Estimating the number of informal caregivers in one region of Switzerland: a population-based study’, Eur. J. Public Health, 2017, [Online]. Available: https://academic.oup.com/eurpub/article/27/suppl_3/ckx189.138/4556920

[31] O. Yip et al., ‘Health and social care of home-dwelling frail older adults in Switzerland: a mixed methods study’, BMC Geriatr., vol. 22, no. 1, Dec. 2022, doi: 10.1186/s12877-022-03552-z.

[32] T. Pan, B. Mulhern, R. Viney, R. Norman, J. Hanmer, and N. Devlin, ‘A Comparison of PROPr and EQ- 5D-5L Value Sets’, Pharmacoeconomics, vol. 40, no. 3, pp. 297–307, Mar. 2022, doi: 10.1007/s40273-021-01109-3.

[33] S. J. Czaja and C. C. Lee, ‘The impact of aging on access to technology’, Univers. Access Inf. Soc., vol. 5, no. 4, pp. 341–349, Mar. 2007, doi: 10.1007/s10209-006-0060-x.

[34] WHO, ‘WHO global strategy on people-centred and integrated health services: interim report’, 2015.

