## Appendix 1 for "The Swiss Integrated Care (INCA) Study: Description of a Novel Prospective Cohort of Patients and Caregivers in Reimbursed Informal Care"

### Details on the Extraction of Diagnostic Information

In addition to structured assessment data, Pflegewegweiser maintains a large repository of clinical documents that are continuously uploaded by patients, informal caregivers, and registered nurses. These uploads comprise both electronically generated documents and scanned files in Portable Document Format (PDF). At the time of upload, each document is assigned to a predefined document category. For the present analysis, only documents classified as medical documents were processed to extract clinical information.

All selected documents were parsed using Azure AI Document Intelligence v3.1, which converts unstructured documents into machine-readable text through advanced Optical Character Recognition (OCR). The extracted text was subsequently processed using Azure OpenAI GPT-4o, a large language model (LLM), following a structured prompt designed to extract diagnoses, medications, administrative information, document metadata, and other clinically relevant medical information. The extracted information from each document was stored in a standardized summary sheet containing the structured outputs of the document analysis.

In a second processing step, all document summary sheets belonging to an individual patient were jointly analyzed using Azure OpenAI GPT-4o to generate a comprehensive patient profile and reconstruct the patient's longitudinal clinical trajectory. By integrating information across multiple documents, the model identified all reported diagnoses, reconciled potentially conflicting information, and assessed the plausibility of each diagnosis. Furthermore, each diagnosis was assigned a temporal classification (acute, chronic, episodic, or resolved) and a confidence level (low, medium, or high), reflecting the model's certainty regarding the validity of the extracted diagnosis. This longitudinal aggregation enables the consolidation of fragmented clinical information into a coherent and structured patient profile suitable for subsequent analyses.

The complete medical document processing workflow is executed within the closed system architecture of Pflegewegweiser. All OCR and LLM components are deployed and executed on a private infrastructure, ensuring that patient documents and extracted information remain within the secure processing environment and are never transmitted to third-party services. Consequently, patient data is not accessible to external providers, including Azure or OpenAI, and cannot be used to train or retrain foundation models. Access to the processing environment is restricted to authorized personnel and authenticated system services only. This architecture ensures compliance with applicable data protection and privacy regulations while maintaining the confidentiality of sensitive patient information.

### Validation of the Extracted Diagnosis Information

Note that the analysis of patients' comorbidity profiles is restricted to chronic diagnoses, with acute and resolved conditions excluded. In contrast, the validation of the information extraction workflow was performed using the complete set of diagnoses to comprehensively assess extraction performance.

The extraction workflow underwent systematic internal validation and quality assessment. For a validation sample of 20 patients, the medical documents were manually reviewed by a medical expert, who identified all confirmed diagnoses reported in the records. This manual review yielded a reference set of 294 diagnoses. Suspected and differential diagnoses were excluded from the reference standard because the objective of the study was to characterize patients confirmed comorbidity profiles. Nevertheless, the extraction pipeline is capable of identifying suspected and differential diagnoses, which are extracted with high precision and assigned exclusively to the 'low'-confidence category, allowing them to be distinguished from confirmed diagnoses while remaining available for applications in which such information is relevant.

Model performance was evaluated by comparing the automatically extracted diagnoses with the manually curated reference standard. Extracted diagnoses were classified as true positives (TP; correctly identified diagnoses), false negatives (FN; missed diagnoses), or false positives (FP; incorrectly extracted diagnoses). False negatives primarily occurred when the LLM failed to recognize a diagnosis within the document. False positives arose when the model incorrectly inferred a diagnosis, for example from a medication list, or generated unsupported diagnoses through hallucinations.

Figure 6 reports the micro-averaged performance metrics used to evaluate the extraction framework:

$$\text{Precision} = \frac{\sum \text{TP}}{\sum \text{TP} + \sum \text{FP}}, \quad \text{Recall} = \frac{\sum \text{TP}}{\sum \text{TP} + \sum \text{FN}}, \quad F_1 = 2\frac{\text{Precision} \times \text{Recall}}{\text{Precision} + \text{Recall}}$$

Performance is evaluated under three scenarios that differ in the confidence categories included in the analysis: (i) only diagnoses classified as high confidence, (ii) diagnoses classified as high or medium confidence, and (iii) all extracted diagnoses (high, medium, and low confidence). Excluding the low confidence category increases precision because this category primarily contains suspected or differential diagnoses. Although these diagnoses are extracted correctly, they are intentionally excluded from the reference standard and therefore count as false positives in the evaluation. At the same time, recall increases slightly when low confidence diagnoses are included, as a small number of confirmed diagnoses are also assigned to this category and would otherwise be counted as false negatives. Restricting the analysis to high confidence diagnoses provides only a marginal gain in precision but leads to a substantial reduction in recall, as diagnoses assigned medium confidence are almost exclusively confirmed diagnoses. Notably, all suspected diagnoses in the validation sample were correctly assigned to the low confidence category by the extraction framework.

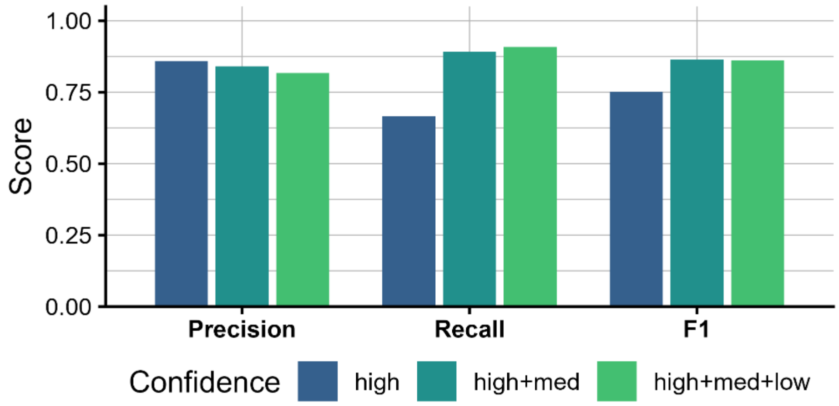


Figure 6: Micro-averaged performance metrics of automatized diagnosis extraction based on a validation sample of 20 patients.

Overall, the combination of high and medium confidence diagnoses achieved the best balance between precision and recall, resulting in the highest performance with an $F_1$-score of 87%. These results demonstrate that the extraction framework is sufficiently accurate and reliable for application in the present study.
